# Incidence of Fractures and Development of the Fracture Risk Model (FRM) in a Population-Based Cohort Study in Abu Dhabi

**DOI:** 10.64898/2026.07.23.26358200

**Authors:** Latifa Baynouna Alketbi, Juma AlKaabi, Shahra Bin Hraiz, Hessa AlNeyadi, Mahra Alyahyaei, Shamsa AlAlawi, Yasmin Mohamed, Maitha Alkaabi, Yahya Alwaqfi, Sara Al Shukri, Shamsa Alshamsi, Sanaa Al Kalbani, Amal Hantash, Ekram Saeed, Waad Alantali, Besent Elbeheiry, AbdulRahman Omara, Amal Al Khouri, Fatima Khalfan AlNuaimi, Mohammad Moussa, Hanan Abdelbaki, Nico Nagelkerke

## Abstract

Fractures in older adults cause high morbidity and mortality. This study aims to identify fracture incidence and risk factors, and to develop the Fracture Risk Model-Abu Dhabi (FRM-AD).

**Method:** This retrospective cohort study of 1757 males and females aged 40+ from Abu Dhabi, United Arab Emirates, who participated in a screening program from 2016 to 2020, followed until 2024 for 4.7 years, SD =1.8. It utilized Electronic Medical Records (EMRs) data and telephone interviews. The outcome assessed was the occurrence of fractures.

**Results:** There were 58 out of the 1149 females (5.0%) and 26 out of the 608 males (4.3%) who had at least one incident fracture. Hip and vertebral fractures accounted for 12.1% and 11.5%, respectively. There was an annual incidence of 10.5 fractures per 1,000 person-years, 10.7 cases among females and 9.9 among males, rising after age 80 to 39.1 among females and 25.3 among males.

Fracture Predictors identified using Poisson regression analysis were older age, smoking, history of previous fracture(s), interaction of history of fracture in a parent with a family history of osteoporosis, positive history of hip fracture in a parent after the age of 40, lack of dyslipidemia diagnosis and the interaction of diabetes mellitus diagnosis and family history of osteoporosis. A higher Body Mass Index (BMI) increased the risk of fractures in this cohort, especially in obese males. Logistic regression of variables at the end of follow-up showed that lower latest vitamin D levels were associated with increased fracture risk.

FRM-AD, derived using Poisson regression, showed moderate discrimination in predicting fractures, with Receiver Operating Characteristic (ROC) curves of 0.731 (0.674-0.788) and 0.742 (0.684-0.800) without and with BMD, respectively. The FRAX AUC to predict MOF and hip fracture ranged between 0.624 and 0.683.

**Conclusion:** In this Emirati cohort, the locally derived FRM-AD showed moderate discrimination for incident fracture and outperformed FRAX-AD, suggesting that a locally derived model may improve fracture risk stratification. Several risk factors and predictors were identified that can be targeted for prevention.

## Introduction

The burden of Major osteoporotic fractures (MOF) is more than that of many noncommunicable diseases, and its incidence is greater than that of cancer, for example. [1] It significantly increases morbidity, hospitalizations, and mortality, and has been described as a global epidemic. Although the global age-standardized rates of fracture incidence, prevalence, and years of healthy life lost (YLDs) decreased slightly between 1990 and 2019, the number of incident cases, prevalent cases, and YLDs increased substantially, largely due to population growth and aging. [2] Fracture risk and its major risk factor, osteoporosis, depend on genetic, metabolic, and environmental interactions. The bone structure in osteoporosis is destroyed to the extent that it becomes fragile and prone to fractures. The complexity of its etiology results in global variability in incidence. This variation in geography, sex, ethnicity, and socio-economic status differs among fracture sites. [3] Key MOF prevention and management instruments are fracture risk assessment tools. These tools are central to clinical guidelines that guide practice and prioritize high-risk patients, and are based on prediction models estimated from available data. The FRAX tool [4] has the most validation studies and has been adapted to diverse ethnicities [5]. Strong evidence suggests that fracture burdens in some countries have been successfully reduced by managing risk factors, including sedentary behavior, smoking rates, diets, and the use of antiosteoporotic medications, as well as increased hip arthroplasties. These positive changes have been linked to efforts by authorities to promote healthy living and access to advanced treatments for risk factors. Therefore, prediction tools, such as the FRAX, are crucial in supporting the expanding screening and management programs and identifying the most susceptible groups. [6], [7].

The application of such assessment predictive tools is complicated by the heterogeneity in risk factors among different settings and the variations in the epidemiology of different types of fractures. [8]. Hip fractures, for example, are the most severe among all MOF fractures in terms of hospitalization, mortality, and patients’ quality of life; [9] and the prediction tools focus mainly on this type of fracture. [10].

Various largely register-based studies have reported substantial worldwide variations in incidence rates, with greater than 10-fold differences between regions [8]. Ethnicity appears to be a major risk factor for MOF, and prediction tools have been designed to accommodate this aspect. The white race has a significantly higher risk than all other races and ethnicities. [11]. As studies on the Arab race(s) are limited, the consensus recommendation is to consider Arabs as white in the use of prediction tools, although there may be unique risk factors, comorbidities, and social factors in the Arab region that may limit their applicability. To date, for example, there is no validation in the Arab population of the FRAX tool, which is the most widely used tool in the Arab regions. As variability in risk factors may affect fracture risk prediction, it would seem necessary to validate this tool for major groups of countries separately. [12].

Specifically, this should take into account variations between regions, where Jordan reports a lower incidence [13] than the Arabian Gulf countries Saudi Arabia, Kuwait, Qatar, and Abu Dhabi, UAE, which showed similarities among each other in the reported incidence of hip fractures. [14] Notably, some studies have shown secular trends in incidence. [15]

In a 2021 review by the GBD 2019 Fracture Collaborators [2], the UAE was listed as a country with a moderate-to-high fracture incidence. Therefore, this study aims to identify the incidence of MOF and its associated risk factors among Emirati adults in Abu Dhabi, United Arab Emirates. It specifically focuses on comparing the data to global trends. Additionally, it seeks to develop a country-specific FRAX model by identifying and highlighting key risk factors and validating the FRAX tool in the Abu Dhabi population.

## Methodology

### Study design

This retrospective cohort study aims to assess the incidence and determinants of fractures among 1757 adult subjects randomly recruited from the community of Abu Dhabi, United Arab Emirates. They included males and females aged 40 years and older. Baseline data, including BMD assessment, were collected for participants in a community-based, free national screening program that began in 2016. The screening, which took place when subjects joined the cohort, was conducted at two primary healthcare centers: one in Abu Dhabi and one in Al Ain, the two largest cities in the Abu Dhabi Emirate. In 2024, an EMR (Electronic Medical Records) chart review was conducted for eligible screening participants, and these subjects were also contacted by telephone to gather further information on lifestyle factors, socio-demographic characteristics, and other potential determinants of fractures.

### Study population

The candidate cohort consisted of 1868 adults aged 40 years and older at the time of screening/ enrollment who participated in a comprehensive preventive program between 2016 and 2020. The program included screening for a wide range of health conditions, including a BMD assessment via dual-energy x-ray absorptiometry (DEXA) at least at one site (hip, spine, or other). Of these 1868 candidates, 17 were unreachable by phone or EMR, and 94 had a DEXA scan performed before or after the included screening period between 2016 and 2020. Therefore, a total of 1,757 subjects were included in the cohort. Appendix 1

### Data collection

First, a data set was retrospectively extracted from the medical records database of participants in the screening program. Data collected included information on participants’ clinical conditions, demographics, lifestyle factors, and laboratory results. Bone mineral density (BMD) was assessed at the time of screening at baseline by dual-energy X-ray absorptiometry (DEXA) of at least one body site: the femoral neck of the hip, vertebral, or other bones. Laboratory results included: Vitamin D levels, HbA1C, liver enzymes, and a lipid profile. Clinical variables obtained from the EMR review included weight and height, blood pressure, medical history, including psychiatric history, cancer history, fatty liver, and CKD history. Data on the use of medications, such as statins, aspirin, osteoporosis medications, vitamin D, and calcium, were also obtained.

In 2024, in addition to the EMR data, telephone interviews were conducted to collect more specific data on lifestyle factors. This phone survey was designed to gather information not typically included in medical records, such as Physical activity (including frequency of walking and resistance training), Occupation, Smoking status, Family history of osteoporosis and fractures above the age of 40, and use of vitamin D and calcium.

To minimize missing data, the included data set from the electronic medical record extracted report included only those with completed baseline data in the screening program. Participants’ information was verified in the EMR cart review and telephone interview. As a result, the proportion of missing values was small, and analyses were conducted on complete cases without imputation.

Two fracture prediction tools were calculated for this cohort of subjects. To validate the tools in this cohort, all necessary variables needed not to be missing. The FRAX (https://www.fraxplus.org/resources) and the Osteoporosis Self-Assessment Tool [16]were used.

### Outcome measure

The primary outcome of this study was the incidence of fractures in the study cohort, defined as fractures diagnosed by a healthcare provider and documented in the electronic medical record with radiological evidence of fracture. Fractures resulting from road traffic collision trauma were excluded as probably not MOF. Secondary outcomes included determinants of fractures, identified by examining the associations of socio-demographic factors, clinical variables, and lifestyle behaviors with fracture risk.

Fractures were ascertained from radiology-confirmed records in the electronic medical record and/or from the telephone interview. Baseline data, including predictor variables, were recorded at or before the baseline screening visit and were collected without knowledge of the fracture outcome.

### Sample size

As a retrospective study, no a priori sample-size calculation was performed; the incidence of fractures in this population was previously unknown, and the maximum number of screening participants with complete, verified data was therefore included. The achieved sample of 1,757 participants, with 84 incident fractures, estimates a fracture incidence of approximately 4.8%, with a 95% confidence interval of ±1.0% (3.8–5.8%), corresponding to the precision attainable from approximately 1,750 participants. For the prediction model, this yielded 84 events; the events-per-variable ratio was adequate.

### Statistical analysis

The Statistical Package for Social Science (SPSS, version 30) was used for the statistical analysis. Descriptive statistics were used to present data. The mean and standard deviation were calculated for continuous variables. Categorical variables were presented as frequencies and percentages. Poisson regression was used to develop a fracture prediction model by examining the variables studied in relation to the occurrence of fractures per unit of time. Candidate predictors were selected a priori on the basis of established fracture risk factors reported in the literature and clinical relevance, rather than by data-driven or stepwise selection; pre-specified interactions were included on the same basis. Logistic regression was used to explore the association between risk factors and “any” fractures (0 = none, 1 = 1+). A *P* value of <0.05 was considered statistically significant. Kaplan-Meier curves and log-rank tests were used to study the time to first fracture. The performance of the fracture models was assessed using the Area Under the Receiver Operating Characteristic (AUC ROC) curve. Determining the optimal cut-off value to predict fracture outcome was through identifying the value at which the ROC curve showed the highest combination of specificity and sensitivity. The FRM-AD score predicts the 10-year cumulative probability of fracture, derived from the Poisson model as 1−exp(−μ) with the linear predictor projected to a 10-year horizon; this horizon matches that of FRAX and enables direct comparison. Internal validation used bootstrap resampling (1,000 replications) to estimate an optimism-corrected area under the ROC curve, the calibration slope, and the calibration-in-the-large intercept, with predicted risk expressed over the cohort’s observed 5-year period. Calibration was additionally assessed using the Hosmer–Lemeshow test, decile-level observed-versus-expected counts, the observed-to-expected event ratio, and flexible (loess) calibration curves, computed with the val.prob function of the rms package in R. Discrimination and the risk cut-offs are reported on the 10-year score, whereas calibration is assessed at the observed 5-year horizon, because absolute risk can only be compared against events actually observed; discrimination is unaffected by the horizon, which is a monotone rescaling.

### Ethical consideration

The study was approved by the Ambulatory Healthcare Services ethics committee, approval number 1-2024. During the calls, all participants provided verbal consent and were informed that their information would be kept confidential and used only for the study. All patient data were anonymized to ensure confidentiality and protect participants’ privacy.

## Results

In this study, the cohort of 1757 subjects, 65.4% were female (1149 individuals), and 34.6% were male (608 individuals). The mean age of participants was 57.95 years (range: 40-93 years; SD = ±9.8), with females averaging 57.1 years (range: 40-93 years; SD = ±9.6) and males averaging 59.6 years (range: 41-88 years; SD = ±9.8). Overall, 3.2% of females and 3.8% of males were 80 or older. At baseline, 40.9% had been diagnosed with diabetes and 28.4% with hypertension. Obesity (defined as a BMI greater than 30 kg/m²) was present in 52.6% of the cohort. Vitamin D levels were adequate (exceeding 75 nmol/L at baseline) among 47.4% of the cohort, while thyroid diagnosis and treatment at baseline were found among 17.1% of the females and 5.6% of the males. At baseline, 83 females (7.2%) and 40 males (6.6%) had fractures before baseline BMD. Additionally, 38 (3.3%) of females and 15 (2.5%) of males reported a parent with a history of fracture after the age of 40 years. Table 1

**Table 1.** Cohort subjects’ baseline characteristics distributed by risk factors studied

|  | Female | Male | Total |
| --- | --- | --- | --- |
| Age groups |  |  |  |
| 40-49 | 299 | 91 | 390 |
|  | 26.00% | 15.00% | 22.20% |
| 50-59 | 469 | 252 | 721 |
|  | 40.80% | 41.40% | 41.00% |
| 60-69 | 260 | 183 | 443 |
|  | 22.60% | 30.10% | 25.20% |
| 70-79 | 84 | 59 | 143 |
|  | 7.30% | 9.70% | 8.10% |
| >=80 | 37 | 23 | 60 |
|  | 3.20% | 3.80% | 3.40% |
| Total | 1149 | 608 | 1757 |
|  | 65.40% | 34.60% | 100.00% |
| Family history of osteoporosis |  |  |  |
|  | 245 | 86 | 331 |
|  | 21.30% | 14.10% | 18.80% |
| Thyroid diagnosis or treatment |  |  |  |
|  | 197 | 34 | 231 |
|  | 17.10% | 5.60% | 13.10% |
| Dyslipidemia before or at screening |  |  |  |
|  | 355 | 177 | 532 |
|  | 30.90% | 29.10% | 30.30% |
| Coronary Artery diseases |  |  |  |
|  | 42 | 29 | 71 |
|  | 3.70% | 4.80% | 4.00% |
| Osteoporosis Before screening |  |  |  |
|  | 106 | 32 | 138 |
|  | 9.20% | 5.30% | 7.90% |
|  | 1149 | 608 | 1757 |
|  | 100.00% | 100.00% | 100.00% |
| Smoking status |  |  |  |
|  | 7 | 107 | 114 |
|  | 0.60% | 17.80% | 6.60% |
| Diabetes Mellitus |  |  |  |
|  | 454 | 265 | 719 |
|  | 39.50% | 43.60% | 40.90% |
| Hypertension |  |  |  |
|  | 27.90% | 29.40% | 28.40% |
| Vitamin D level |  |  |  |
| <30 | 24 | 13 | 37 |
|  | 2.20% | 2.40% | 2.30% |
| 31-49 | 208 | 92 | 300 |
|  | 19.00% | 16.80% | 18.20% |
| 50-74 | 364 | 165 | 529 |
|  | 33.20% | 30.20% | 32.20% |
| 75-149 | 501 | 276 | 777 |
|  | 45.70% | 50.50% | 47.30% |
| 150-299 | 0 | 1 | 1 |
|  | 0.00% | 0.20% | 0.10% |
| Cancer |  |  |  |
|  | 43 | 12 | 55 |
|  | 3.70% | 2.00% | 3.10% |
| Fracture Before screening BMD |  |  |  |
|  | 83 | 40 | 123 |
|  | 7.20% | 6.60% | 7.00% |
| History of fracture in the famiyy |  |  |  |
|  | 143 | 57 | 200 |
|  | 12.40% | 9.40% | 11.40% |
| History of fracture in a parent after the age of 40 |  |  |  |
|  | 38 | 15 | 53 |
|  | 3.30% | 2.50% | 3.00% |
| Bone health at screening BMD |  |  |  |
| Normal BMD | 527 | 343 | 870 |
|  | 45.90% | 56.40% | 49.50% |
| Osteopenia | 383 | 171 | 554 |
|  | 33.30% | 28.10% | 31.50% |
| Osteoprosis | 239 | 94 | 333 |
|  | 20.80% | 15.50% | 19.00% |
| Obesity classes |  |  |  |
| <25 | 108 | 126 | 234 |
|  | 9.40% | 20.70% | 13.30% |
| 25-29.999 | 333 | 267 | 600 |
|  | 29.00% | 43.90% | 34.10% |
| 30-34.999 | 403 | 155 | 558 |
|  | 35.10% | 25.50% | 31.80% |
| >=35 | 305 | 60 | 365 |
|  | 26.50% | 9.90% | 20.80% |
| Total | 1149 | 608 | 1757 |

B. Incidence of fracture during follow-up
| Fracture after screening BMD |  |  |  |
| --- | --- | --- | --- |
|  | 58 | 26 | 84 |
|  | 5.00% | 4.30% | 4.80% |
| No fracture | 1091 | 582 | 1673 |
|  | 95.00% | 95.70% | 95.20% |
| One fracture | 50 | 24 | 74 |
|  | 4.40% | 3.90% | 4.20% |
| Two fractures | 8 | 2 | 10 |
|  | 0.70% | 0.30% | 0.60% |

Regarding the use of medications and supplements at the end of follow-up among subjects who experienced at least one fracture, 32.8% of females and 30.7% of males were on anti-resorptive medications, compared to 26% of females and only 13.8% of males in the group with no fractures. This cohort reported high use of vitamin D supplementation, with 21.2% and 24.4% of individuals not using any in the fracture and non-fracture groups, respectively. Adherence was similar in both groups, with high use rates of 78.8% and 71.4% in the fracture and non-fracture groups, respectively. The irregular use of vitamin D was 4.2% in the non-fracture group and none in the fracture group. Regarding calcium use, 54.8% of those with fractures were on it compared to 37.3% without fractures. The combination of vitamin D and calcium was used by only 38.1% of those with fractures and 27.2% of those without. Males with no fracture used a combination less often than females, 19.1% compared to 31.5%. Statin use was common in this cohort, with 76.9% of those with fractures and 69.3% without fractures, while Aspirin use was 31% and 21.5%, respectively.

### Incidence of fracture

At the end of the follow-up period (averaging 4.7 years, SD =1.8, range 0 – 8), 58 out of 1149 females (5.0%) and 26 out of 608 males (4.3%) had at least one incident fracture following the baseline BMD assessment at the beginning of the follow-up. There were 10 subjects (11.9% of those with a fracture), consisting of eight females (13.8% of females with a fracture) and two males (7.7% of males with a fracture), who experienced two fractures. From ages 40 to 60, the cumulative incidence of fractures over the follow-up period is about 3%, roughly doubling between ages 60 and 80 (6.5% and 6.3%), and doubling again after age 80 (15.0%). Figure 1 illustrates the difference in incidence between males and females across age groups.

**Figure 1.**
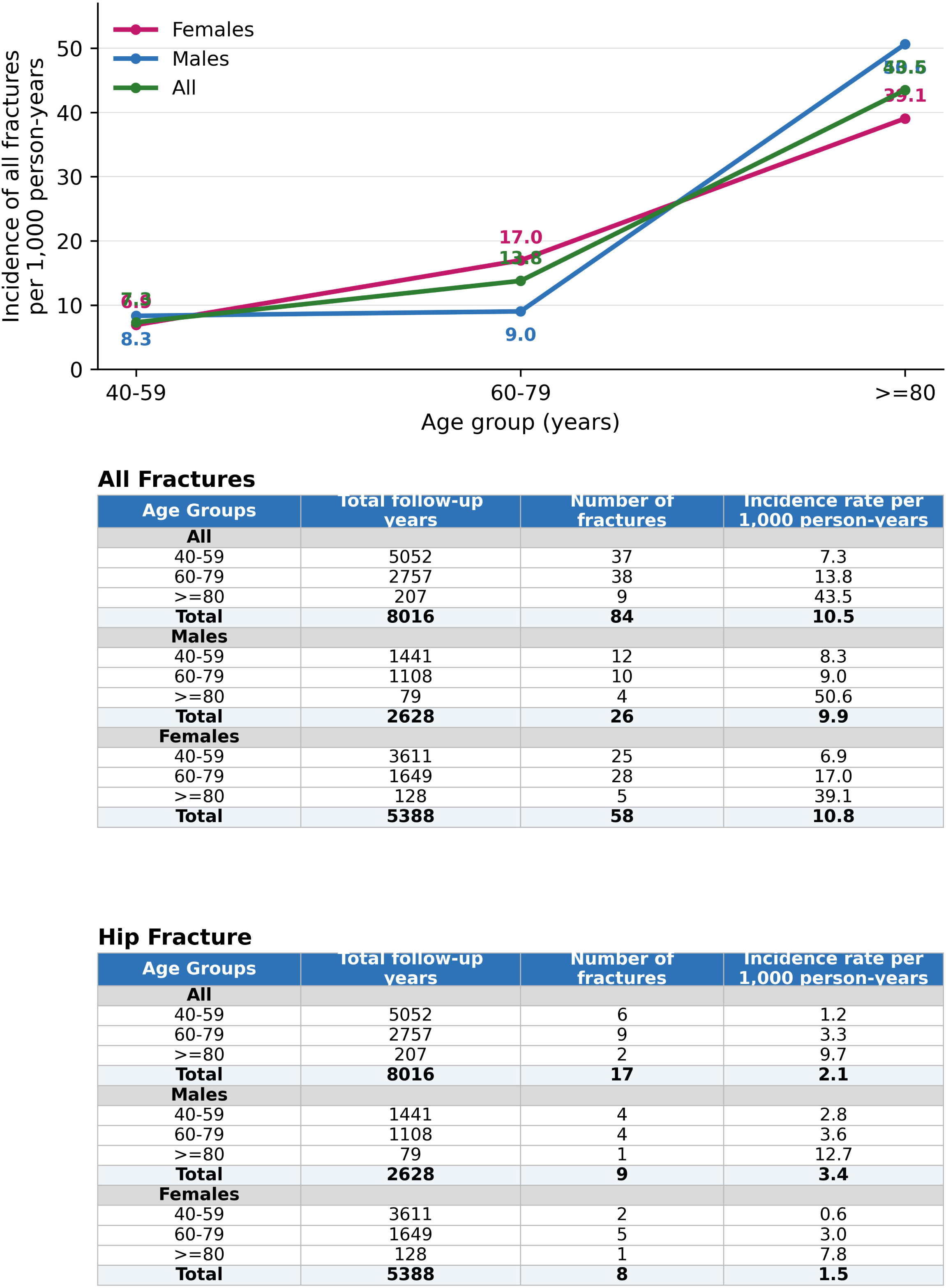
Annual incidence of fracture

Among this cohort, the first fractures following the baseline BMD assessment were hip fractures in 12.1%, vertebral fractures in 11.5%, foot and toe fractures in 25.5%, hand and arm fractures in 27.4%, leg fractures in 12.7%, and wrist fractures in 6.7%. Vertebral and hip fractures accounted for 23.6% of all subsequent fractures.

The incidence of fracture per 1,000 person-years annually was 10.7 cases among females and 9.9 among males, resulting in a female-to-male ratio of 1.1. Females had a higher incidence in their 60s to 70s, at 17.0 per 1,000 person-years, compared with 10.8. However, the incidence of fractures increased substantially among both sexes at ages 80 and older, with females having a higher incidence of 39.1 per 1,000 person-years, compared to 25.3 among males. Based on 17 events, the incidence of hip fracture was 2.1 per 1,000 person-years (95% CI 1.3-3.4). Figure 1

### Fracture risk factors

Although the female-to-male ratio is variable and depends on the fracture site and age, Poisson regression analysis performed to assess predictors of fracture (all sites combined) showed no significant difference between females and males, Table 2 A and B, without BMD included and with BMD, p-value 0.118 without BMD and p=0.241 when BMD is added. Older age was associated with higher fracture risk; for each year older, the risk increased by 3.2%, exp(0.032)=1.032, P=0.004, and by 2.4% for every year when the BMD is added as a variable, exp(0.024)= 1.024 (p-value =0.043), that is increased risk of 37% and 27% for each decade of age without and with BMD respectively. Smoking was a marginally significant predictor of fracture in the model without BMD (p = 0.071). However, when BMD was included, smoking was a significant predictor: non-smokers had a 63% lower risk (exp(-0.997) = 0.37; equivalently, smokers had an approximately 2.7-fold higher risk), p = 0.02. Regarding the reporting history of fractures before, in the model without BMD the absence of a prior fracture was associated with 43% lower risk (exp(-0.564) = 0.57); equivalently, a prior fracture trended toward higher risk, but this was only marginally significant (p = 0.074). Table 2 A and B The interaction of history of fracture in a parent with family history of osteoporosis was significant predictor in both models with and without BMD with subjects with negative family history of osteoporosis and not having or having history of fracture in a parent above 40years are protected more than other subjects, exp(-1.605)=0.2, P value 0.005, and exp(-1.812)=0.163 (p-value 0.005), respectively. The same significant interaction was observed when adding BMD: exp(-1.399) = 0.25 (p-value 0.015) and exp(-1.798) = 0.165 (p-value 0.006), respectively. Worth noting that subjects with a positive history of fracture in a parent, having a negative family history of osteoporosis, do not place them at lower risk.

**Table 2.** Predictors of fractures in 1757 subjects. The dependent variable is the number of fractures using Poisson regression A. With no BMD included. B. With BMD included

| Parameter | B | 95% Wald Confidence Interval |  | P value |
| --- | --- | --- | --- | --- |
|  |  | Lower | Upper |  |
| (Intercept) | -4.332 | -6.183 | -2.48 | <.001 |
| Female | 0.423 | -0.107 | 0.954 | 0.118 |
| Male | 0<sup>a</sup> | . | . | . |
| Not Smoker | -0.761 | -1.587 | 0.064 | 0.071 |
| Smoker | 0<sup>a</sup> | . | . | . |
| No history of fracture before screening | -0.564 | -1.182 | 0.055 | 0.074 |
| History of fracture before screening | 0<sup>a</sup> | . | . | . |
| Age | 0.032 | 0.01 | 0.053 | 0.004 |
| No Dyslipidemia | 0.452 | 0.021 | 0.882 | 0.04 |
| Dyslipidemia | 0<sup>a</sup> | . | . | . |
| No Diabetes Mellitus | -1.111 | -2.478 | 0.256 | 0.111 |
| Diabetes Mellitus | 0<sup>a</sup> | . | . | . |
| BMI deviation from this cohort mean | 0.302 | 0.082 | 0.523 | 0.007 |
| The interaction of no history of hip fracture of a parent after the age of 40 and no family history of osteoporosis | -1.605 | -2.723 | -0.487 | 0.005 |
| The interaction of no history of hip fracture of a parent after the age of 40 and family history of osteoporosis | -1.812 | -3.089 | -0.535 | 0.005 |
| The interaction of history of hip fracture of a parent after the age of 40 and no family history of osteoporosis | -1.039 | -2.619 | 0.541 | 0.197 |
| The interaction of history of hip fracture of a parent after the age of 40 and family history of osteoporosis | 0<sup>a</sup> | . | . | . |
| No history of hip fracture of a parent after the age of 40 | 0<sup>a</sup> | . | . | . |
| history of hip fracture of a parent after the age of 40 | 0<sup>a</sup> | . | . | . |
| No family history of osteoporosis | 0<sup>a</sup> | . | . | . |
| Family history of osteoporosis | 0<sup>a</sup> | . | . | . |
| The interaction of no Diabetes and no family history of osteoporosis | 1.331 | -0.105 | 2.767 | 0.069 |
| The interaction of Diabetes and no family history of osteoporosis | 0<sup>a</sup> | . | . | . |
| The interaction of no Diabetes and family history of osteoporosis | 0<sup>a</sup> | . | . | . |
| The interaction of Diabetes and family history of osteoporosis | 0<sup>a</sup> | . | . | . |
| Vitamin D deviation from this cohort mean | 0.002 | -0.004 | 0.008 | 0.563 |
| The interaction of female sex and BMI deviation from this cohort mean | -0.134 | -0.213 | -0.054 | 0.001 |
| The interaction of male sex and BMI deviation from this cohort mean | 0<sup>a</sup> | . | . | . |
| The interaction of age and BMI deviation from this cohort mean | -0.003 | -0.007 | -1.47E-05 | 0.049 |
| (Scale) | 1<sup>b</sup> |  |  |  |

B. With BMD included
| Parameter | B | 95% Wald Confidence Interval |  | P value |
| --- | --- | --- | --- | --- |
| (Intercept) | -3.274 | -5.269 | -1.278 | 0.001 |
| Female | 0.327 | -0.219 | 0.872 | 0.241 |
| Male | 0<sup>a</sup> | . | . | . |
| Not Smoker | -0.997 | -1.837 | -0.157 | 0.02 |
| Smoker | 0<sup>a</sup> | . | . | . |
| No history of fracture before screening | -0.409 | -1.059 | 0.241 | 0.218 |
| History of fracture before screening | 0<sup>a</sup> | . | . | . |
| Age | 0.024 | 0.001 | 0.047 | 0.043 |
| No Dyslipidemia | 0.52 | 0.079 | 0.961 | 0.021 |
| Dyslipidemia | 0<sup>a</sup> | . | . | . |
| No Diabetes Mellitus | -0.987 | -2.36 | 0.386 | 0.159 |
| Diabetes Mellitus | 0<sup>a</sup> | . | . | . |
| BMI deviation from this cohort mean | 0.099 | 0.034 | 0.165 | 0.003 |
| The interaction of no history of hip fracture of a parent after the age of 40 and no family history of osteoporosis | -1.399 | -2.527 | -0.27 | 0.015 |
| The interaction of no history of hip fracture of a parent after the age of 40 and family history of osteoporosis | -1.798 | -3.085 | -0.512 | 0.006 |
| The interaction of history of hip fracture of a parent after the age of 40 and no family history of osteoporosis | -0.923 | -2.505 | 0.658 | 0.253 |
| The interaction of history of hip fracture of a parent after the age of 40 and family history of osteoporosis | 0<sup>a</sup> | . | . | . |
| No history of hip fracture of a parent after the age of 40 | 0<sup>a</sup> | . | . | . |
| history of hip fracture of a parent after the age of 40 | 0<sup>a</sup> | . | . | . |
| No family history of osteoporosis | 0<sup>a</sup> | . | . | . |
| Family history of osteoporosis | 0<sup>a</sup> | . | . | . |
| The interaction of no Diabetes and no family history of osteoporosis | 1.25 | -0.2 | 2.7 | 0.091 |
| The interaction of Diabetes and no family history of osteoporosis | 0<sup>a</sup> | . | . | . |
| The interaction of no Diabetes and family history of osteoporosis | 0<sup>a</sup> | . | . | . |
| The interaction of Diabetes and family history of osteoporosis | 0<sup>a</sup> | . | . | . |
| Vitamin D deviation from this cohort mean | 0.003 | -0.004 | 0.009 | 0.438 |
| The interaction of female sex and BMI deviation from this cohort mean | -0.136 | -0.215 | -0.056 | <.001 |
| The interaction of male sex and BMI deviation from this cohort mean | 0<sup>a</sup> | . | . | . |
| Normal BMD | -1.018 | -1.613 | -0.423 | <.001 |
| Osteopenia | -0.647 | -1.191 | -0.103 | 0.02 |
| Osteoporosis | 0<sup>a</sup> | . | . | . |
| Thyroid Stimulating Hormone deviation from this cohort mean | 0.003 | -0.045 | 0.052 | 0.898 |
| (Scale) | 1<sup>b</sup> |  |  |  |

Regarding Body Mass Index, a higher BMI was associated with an increased fracture risk in this cohort, even after controlling for the significant interactions between sex and BMI and between age and BMI. This indicates a higher fracture risk among obese older individuals and obese males. This was evident in both analyses, with and without BMD. For each 1-unit increase in BMI above the cohort mean, the fracture risk increases by 35% (exp(0.302) = 1.35), P=0.007. Kaplan-Meier plots for time to first fracture, shown in Figure 2, revealed that higher BMI classes were associated with a greater fracture incidence in males and a lower incidence in females, although the log-rank test was not significant (P=0.4).

**Figure 2.**
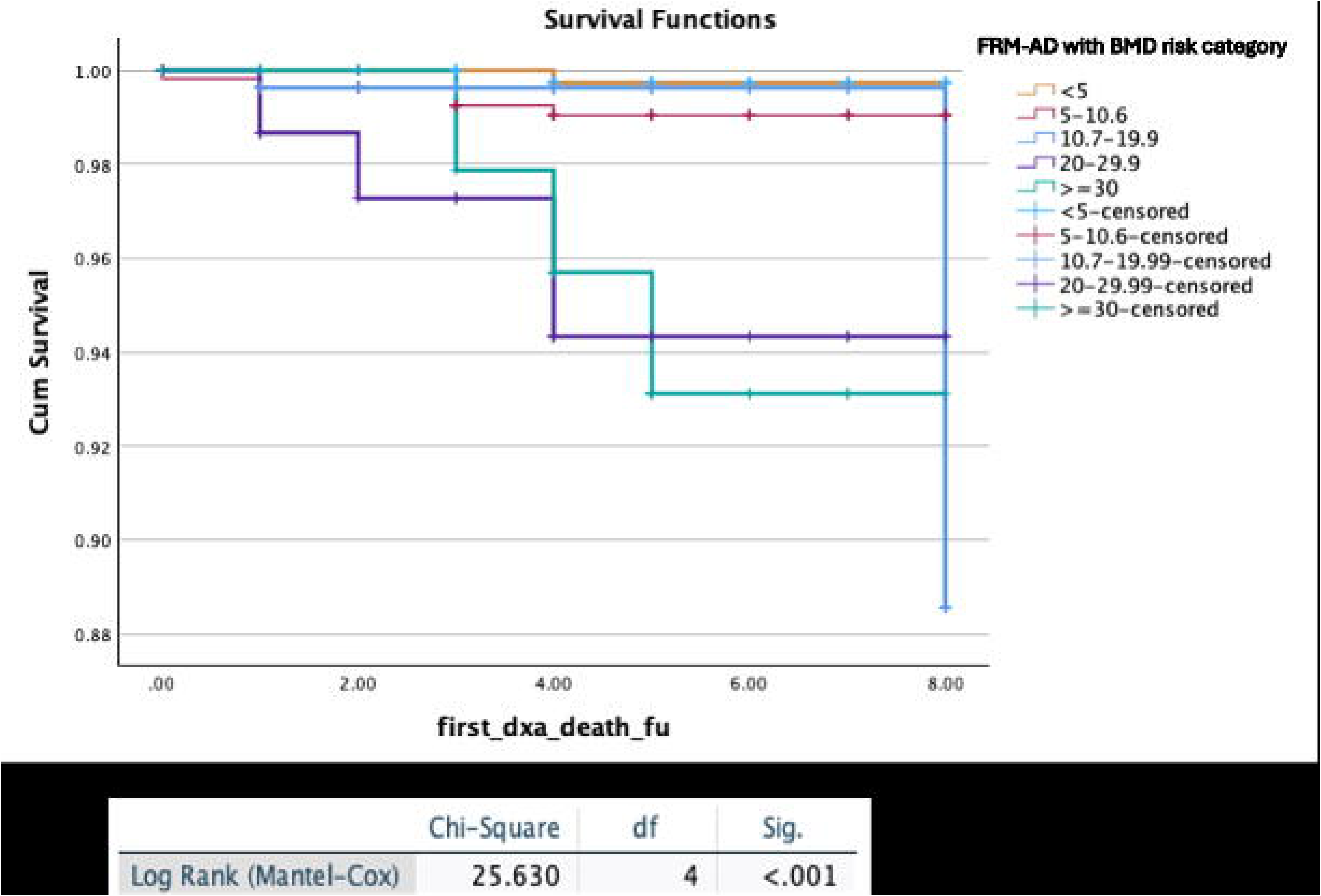
Kaplan-Meier plots for the time to first fracture as per BMI classes among males and females

Several interesting risk factors were identified in this cohort. First was the interaction of diabetes mellitus diagnosis and family history of osteoporosis. Subjects without diabetes and who are not reporting a family history of osteoarthritis did have a higher risk of fracture. On the other hand, a lack of a dyslipidemia diagnosis placed subjects at a significantly higher risk of fracture in both analyses, with and without BMD: exp(0.452) = 1.57, P = 0.04; exp(0.52) = 1.68, P = 0.021, respectively.

Meanwhile, other risk factors studied, such as baseline vitamin D levels, showed no significant associations. A diagnosis of thyroid disease and a history of fracture before screening were not significant predictors, whether or not they included BMD.

### Derived FRM-AD

Both models derived from Poisson regression, one without BMD and one with BMD, showed moderate discrimination in predicting fractures. The Receiver Operating Characteristic (ROC) curve is illustrated in Table 3, Appendix 2. The model developed without BMD achieved an AUC of 0.731 (0.674-0.788), a sensitivity of 69.6%, and a specificity of 73.9% at a cut-off risk of 10.8, indicating moderate discrimination between individuals with and without fracture. The model with BMD performed similarly, with an AUC of 0.742 (0.684-0.8), a sensitivity of 71.1%, and a specificity of 71.5% at a cut-off point of risk of 10.6. Table 3, A On internal validation by bootstrapping (1,000 replications), discrimination was good with minimal optimism: the apparent AUC was 0.731 without BMD and 0.742 with BMD, and fell only marginally after bias correction, to optimism-corrected values of 0.73 and 0.74, respectively (a difference of less than 0.01), indicating that the model is not overfit. When predicted risk was expressed over the cohort’s observed 5-year horizon, calibration was good in both models. The calibration slope was 1.01 in both, and the calibration-in-the-large intercept was −0.03 without BMD and −0.04 with BMD; the joint test of calibration intercept and slope was non-significant (χ² = 0.09, p = 0.96 without BMD; χ² = 0.08, p = 0.96 with BMD). The observed-to-expected event ratio was 0.97 (95% CI 0.78–1.21; 79 observed versus 81.5 expected events) without BMD and 0.97 (95% CI 0.77–1.21; 76 versus 78.3 events) with BMD, and the Hosmer–Lemeshow test was non-significant in both (χ² = 13.50, df = 8, p = 0.10 without BMD; χ² = 10.29, df = 8, p = 0.25 with BMD). The average absolute calibration error was 0.8% without BMD and 0.7% with BMD, with 90th-percentile absolute errors of 1.6% and 1.4% respectively. The flexible calibration curve lay close to the ideal line across the range containing the large majority of participants, with modest under-prediction at intermediate predicted risk (Figure 3). Few participants had high predicted risk, only 51 had a predicted 5-year risk above 15% and 20 above 20%,so the nonparametric curve became unstable and diverged from the diagonal in the upper range (maximum absolute error 0.51 without BMD and 0.23 with BMD), and calibration in that region could not be reliably assessed. External validation in a larger, independent cohort is nonetheless warranted before clinical use. A larger cohort would also include more individuals at high predicted risk, enabling calibration to be verified across the full range of risk levels—including the high-risk range that the present sample was too small to evaluate.

**Figure 3.**
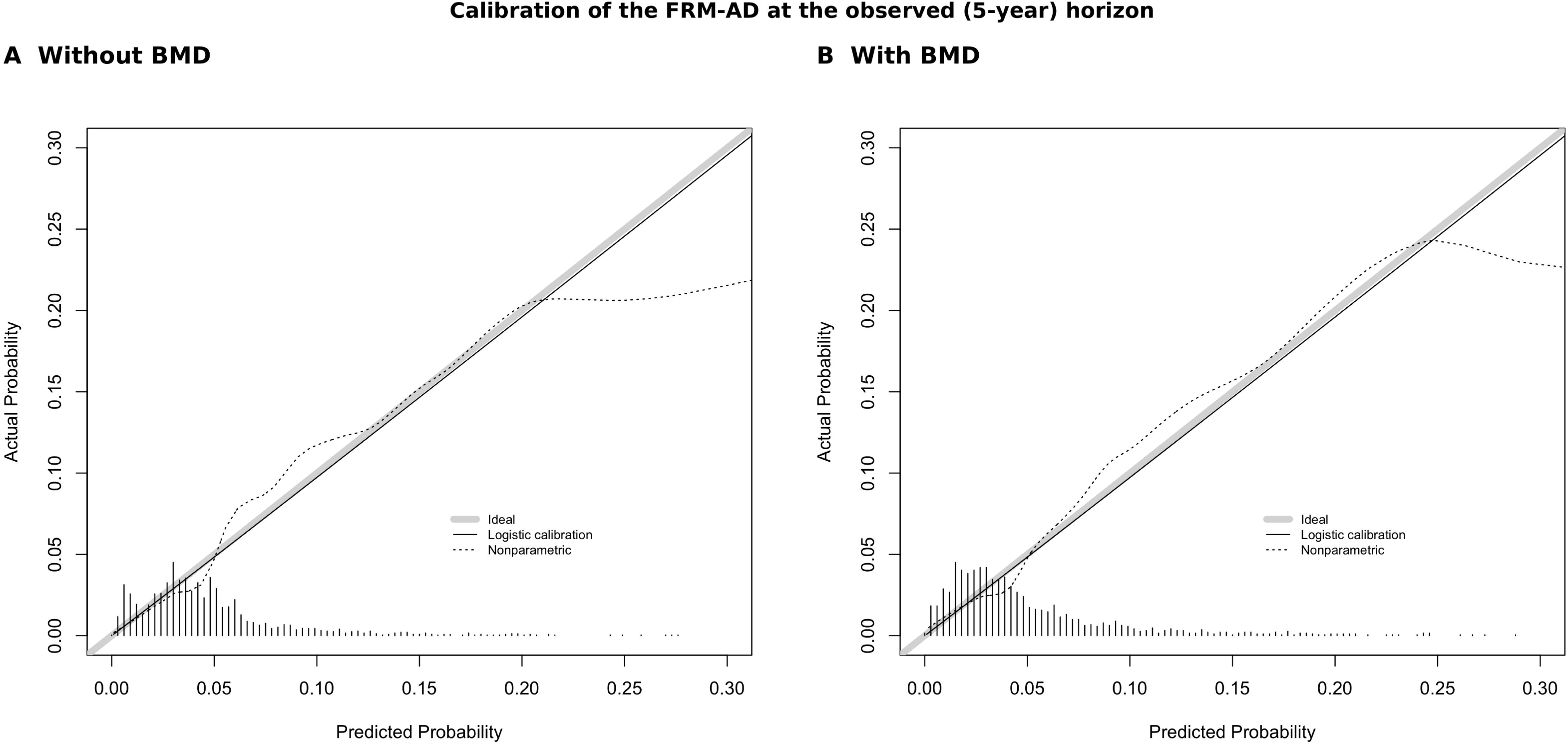
Calibration of the FRM-AD at the observed (5-year) horizon. (A) Without BMD; (B) with BMD. The grey line denotes ideal calibration, the solid line the logistic-calibration estimate, and the dashed line the nonparametric (loess) estimate; tick marks show the distribution of predicted risk.

**Table 3.** (A)The performance of the derived model to predict the 10-year probability of fracture. (B) The performance of the FRAX-Abu Dhabi and Osteoporosis Self-Assessment Tool (OST) predicts the 10-year probability of fracture (C). The performance of the age-only, hip BMD-only, and vertebral-only models in predicting the 10-year probability of fracture.

|  |  | n | Fractures | AUC | 95% CI | Cutoff | Sensitivity | Specificity |
| --- | --- | --- | --- | --- | --- | --- | --- | --- |
| FRM-AD-no BMD (MOF) | Both sex | 1,645 | 79 | 0.731 | 0.674-0.788 | 10.8 | 69.6 | 73.9 |
|  | Female | 1,098 | 56 | 0.751 | 0.687-0.815 | 10.8 | 75.0 | 71.8 |
|  | Male | 547 | 23 | 0.677 | 0.558-0.796 | 12.1 | 56.5 | 82.4 |
| FRM-AD BMD (MOF) | Both sex | 1,509 | 76 | 0.742 | 0.684-0.801 | 10.6 | 71.1 | 71.5 |
|  | Female | 1,022 | 54 | 0.760 | 0.691-0.828 | 11.8 | 72.2 | 74.4 |
|  | Male | 487 | 22 | 0.703 | 0.593-0.813 | 9.7 | 63.6 | 73.1 |

The model derived from this study was used to predict hip fractures. Although the number of these fractures was low, eight females and nine males, the AUC was satisfactory. For the model with BMD, the AUC was 0.723 (0.599-0.846), and for the one without BMD, it was 0.722 (0.584-0.861). The models performed better among females than males. AUC difference between females and males was 0.268 (0.878 compared to 0.610) in the model with BMD and 0.256 (0.874 compared to 0.618) in the model without BMD.

The performance of the model at different cutoff points is shown in Appendix 5, indicating the best performance at 10, with an excellent negative predictive value and a positive predictive value of 10.1 without BMD and 10.8 with BMD.

### Risk factors identified from logistic regression, with risk factors reported at the end of follow-up

Although vitamin D at baseline was not a significant determinant of fractures in Poisson regression, in the analysis using logistic regression with the incidence of at least one fracture after baseline BMD and the latest values of risk factors at the end of follow-up as covariates, a lower vitamin D level was associated with higher fracture incidence. The likelihood of having a fracture increased by 16% for each 10-unit reduction in vitamin D level, OR = 0.984 (0.972-0.996), p = 0.009. Age, sex, and BMI were not significantly associated, but lower LDL, smoking, not having dyslipidemia before, a history of hip fracture in a parent, and a diagnosis of osteoporosis were.

### FRAX-AD validation on this cohort and in comparison with the FRM-AD

Of the 1757 subjects, 275 had at least one missing FRAX variable, leaving 1482 for FRAX validation. The Fracture Risk Assessment Tool (FRAX), both without and with bone mineral density added, performed acceptably for MOF and hip fracture prediction. The AUC for FRAX performance without BMD to predict MOF was 0.624 (0.545-0.703), and 0.623 (0.548-0.698) with BMD. The FRAX-MOF AUC among males is 0.466/0.471 with 95% CI (0.31-0.62), but there were few male events. Table 3

For Hip fracture prediction, the AUC for FRAX performance without BMD was 0.638(0.479-0.796). When the BMD was included, the FRAX performed much better with an AUC of 0.683(0.571-0.795). Similarly, the performance of the FRAX was a better predictor of hip fracture among females than males. Table 3 The FRAX with BMD included performed best in predicting hip fractures among females, with an AUC reaching 0.754 (0.661-0.847).

Worth noting that when applying the cutoff point of the FRAX, which is 20% for MOF and 3% for hip fractures, the sensitivity was very low, but the specificity was excellent. For FRAX (MOF) without and with BMD, the sensitivity was approximately 1%, and the specificity was 99.9%. For predicting hip fracture at the 3% cutoff, the sensitivity of FRAX (MOF) was around 10% with and without BMD, and the specificities were 97.6% and 93.5%, respectively. At the cutoff points identified by ROC analysis, as shown in Table 3, with a cutoff around 2.8 for MOF and 0.25 for hip, the sensitivity was between 54% and 58.7% for predicting MOF and 80% for predicting hip, respectively. The specificity ranged from 68.4% to 74.2% for MOF and around 60% for hip.

The Osteoporosis Self-Assessment Tool (OST) performed slightly better than FRAX, with an AUC of 0.63 (0.562-0.697). The AUC was 0.670 (0.599-0.742) among females and 0.545 (0.403-0.686) among males. This is close to using age to predict fracture incidence, which showed an AUC of 0.664 (0.595-0.734) among females and 0.586 (0.458-0.715) among males. Or using the BMD only, which yielded an AUC of 0.641 (0.564-0.719) at best among females using the vertebral T-score, but it was poor among males. At best, for males, the AUC was 0.578 (0.465-0.691) using the hip T-score. Table 3, Appendix 2

### Applying the FRM-AD

Over an average of 4.7 years, at the ≥10% cutoff, the FRM-AD identified as high-risk about 70% of those who subsequently fractured (single-model sensitivity 69.6% at the 10% cutoff without BMD; Appendix 5). Because incident fractures were uncommon, only about 10% of high-risk individuals actually fractured (PPV 10%), whereas 98% of those classified as low risk remained fracture-free (NPV 98%). In contrast, one in five of those with two fractures would have been missed because they had a low-risk score. Among those with one fracture, 35.4% of females and 47.6% of males were categorized as low risk according to the FRM-AD without BMD, and 23.9% of females and 45% of males were categorized as low risk according to the FRM-AD with BMD. Overall, 74.6% of those who had at least one fracture were identified either with the FRM-AD with BMD or FRM-AD without BMD as high risk. As shown in Appendix 3, when both FRM-AD with and without BMD were used, 61.9% of males and 79.2% of females who developed one fracture, and 8 of the 10 subjects who developed two fractures, were classified as high risk (FRM-AD was >= 10%). This could improve with a longer follow-up period, as the score was calculated based on a 10-year risk. Time to first fracture stratified by FRM-AD (with BMD) risk category is shown in Appendix 4; fracture-free survival separated significantly across risk categories (log-rank P < 0.001).

## Discussion

The all-fracture incidence in this study (10.5 per 1,000 person-years) is comparable to rates reported in several high-income countries. The incidence of fractures ranged from 10.9 per 1,000 person-years in Spain to 11.6 per 1,000 person-years in the United Kingdom and 12.29 per 1,000 person-years in Sweden. In neighboring countries, there are no reports of total fracture incidence, but the hip fracture incidence in this study was 2.1 per 1000 person-years annually, close to the 1.0 per 1000 person-years annually reported in Saudi Arabia among females and 0.7 per 1000 person-years annually among males. In Kuwait and Iran, it was similar between the two sexes, with 1.5 among females and males. [17]

This study identified several significant predictors of fractures among adults aged 40 and above in the United Arab Emirates (UAE). Shared risk factors with other published fracture prediction tools are age, smoking, a parent’s history of hip fracture, and osteoporosis. But other identified risk factors highlight areas for interventions to modify risk and prevent fractures. The near-absence of a sex difference in overall fracture risk is notable and warrants further investigation. In most populations women carry a substantially higher fracture burden, in the US, for example, about 71% of fractures occur in women, whereas in this cohort the all-fracture incidence was similar in the two sexes (10.7 per 1,000 person-years in women and 9.9 in men). The pattern was site-and age-dependent: women had higher all-fracture rates at the oldest ages, but men had a higher hip-fracture incidence (3.4 versus 1.5 per 1,000 person-years) and higher all-fracture rates in the 70–79 age band, with nearly one in twenty men sustaining a fracture overall. The inclusion of minor fractures and the small number of hip fractures, together with wide confidence intervals in the oldest age groups, limit firm conclusions about age- and sex-specific patterns. Nonetheless, the comparatively high fracture burden in men, and particularly their higher hip-fracture rate, suggests that men in this population may be more vulnerable than the conventional focus on women would imply. Behavioral and social factors, such as greater outdoor activity among older men in Abu Dhabi, may contribute and warrant further study. Notably, the higher prevalence of osteoporosis among women than men in this same population [17]did not translate into a correspondingly higher fracture incidence in women, underscoring that bone mineral density alone does not fully capture fracture risk in this setting.

This pattern aligns with secular trends observed in Western populations, where the historically large female-to-male fracture gap has been narrowing. Age-standardized hip fracture rates have declined more rapidly in women than in men, with male rates in some cohorts remaining stable or even rising, particularly among the oldest old [43, 44, 45]. A recent multinational analysis similarly reported smaller declines in incidence among men, a substantial post-fracture treatment gap in men, and a projected increase in the number of hip fractures by 2050 that is greater in men than in women [46]. Against this backdrop, the absence of a significant sex difference and the high incidence among men aged 80 and older in this cohort may reflect a broader epidemiological shift rather than an isolated finding, reinforcing the argument that fracture prevention and osteoporosis screening should target both sexes. The USPSTF’s current recommendation is that evidence is insufficient to assess the balance of benefits and harms of screening for osteoporosis to prevent osteoporotic fractures in men [18]. At least in Abu Dhabi, this study suggests that both sexes should be treated equally in screening. Another possible explanation for the differences in fracture risk between men and women is that they may stem not only from differences in areal BMD but also from variations in bone size, geometry, and strength. In Abu Dhabi, the prevalence of osteoporosis is among the highest in the world, 22.9%, 25.1% among women, and 18.8% among men, with only a female-to-male ratio of 1.3, indicating an added risk to Abu Dhabi men compared to other countries. [17] Worldwide, the prevalence of osteoporosis is reported to be more than double in women, 23.1%, compared with 11.7% in men [17] [19]. Additionally, it is reported that males with OP are less likely to be on anti-resorption treatment than females [20], and this can add to fracture risk. There is no data on baseline OP treatment in this cohort, but at follow-up, there was no difference in treatment between the two sexes in the group with fracture, unlike the group with no fracture, which showed double the likelihood of being treated with anti-resorptive treatment and vitamin D and calcium combination among females, which supports the undertreatment hypothesis.

Unsurprisingly, age was a significant factor, with a 3.2% increase in risk for every year of age, remaining at 2.4% after adjusting for BMD. Nevertheless, the influence of BMI on fracture risk also shows different expressions. Obesity is a greater risk factor for fracture. The effect of BMI on the risk of fractures in our cohort interacted with sex, which was similar to other studies. Obesity was found to be associated with fracture, which at least partially may be explained by worse physical function in obese men. [21], [22], [23]. A study from Australia concluded that the observed positive association between BMI and fracture risk is mediated by femoral neck BMD in both men and women [24] [25] [26]. On the other hand, body fat composition is variable even with normal BMI, and lean mass exerts a greater effect on BMD than fat mass in men and women combined, emphasizing the role of physical activity in bone health. [27], [28]

The risk of fracture decreases with a diagnosis of dyslipidemia, regardless of the use of statins, which is difficult to interpret, especially since hypercholesterolemia was found to be associated with reduced BMD, and ex vivo, in vivo, and clinical studies suggest that lipid-lowering agents reduce both atherosclerotic calcification and osteoporosis [29]. Nevertheless, non-linear U-shaped relationships with hip fracture risk were described. A recent study investigated the associations between concentrations of lipid metabolites and osteoporotic fractures in both men and women. There were 35 exhibited significant associations with incident fractures in patients diagnosed with osteoporosis, with male patients having an increased risk, and some have been shown to decrease the risk of incident fractures in female patients. The precise mechanisms through which these molecules contribute to fractures in osteoporosis remain unclear. The authors stated that studies on the roles of lipids in the development of osteoporosis are in their early stages and that more research will be necessary. [30]

The higher risk of fracture among subjects with no family history of osteoporosis and no history of diabetes may be attributed to health behaviors, including fewer visits to healthcare providers, which lead to less health promotion and risk factor management. Similarly, the increased risk among those with a family history of osteoporosis who have no history of hip fracture in a parent above 40 may be attributed to awareness and early implementation of preventive measures.

The Poisson regression model was comparable in performance to the better-performing fracture assessment tools reported in the literature. [5] [31] This finding may have implications for clinical practice. Healthcare providers should prioritize osteoporosis screening in individuals with multiple risk factors and, where appropriate, initiate therapeutic options in accordance with treatment guidelines. Implementing this fracture risk score and personalized preventive strategies could significantly reduce the incidence of fractures. In particular, fall prevention effectively reduces the number of fallers and fall rate in a network meta-analysis by et al. [32],[33]. Such prevention encompasses exercise as a single intervention, assistive technology, environmental assessment and modifications, and basic fall risk assessment (e.g., medication review).

The FRAX tool, the most widely used assessment in patient care, has been shown to perform well in fracture prediction across many studies. The FRAX-AD was adapted using local population data. Because both tools express the 10-year probability of fracture, the FRM-AD and FRAX-AD were compared on discrimination, which is horizon-matched; the absolute calibration of FRAX-AD could not be assessed in this cohort because its 10-year horizon exceeds the observed follow-up. Among women, its performance in this cohort was similar to the findings reported in the US studies, the Study of Osteoporotic Fractures (SOF) cohort, and the Women’s Health Initiative cohort (AUC 0.64 in SOF versus 0.64 in WHI). [34] The improved performance of this study’s developed tool, FRM-AD, may reflect changes in the local population over time, including lifestyle choices, medication use, changes in risk factor trends, and variations in its interactions. This emphasizes the necessity of considering these factors in locally developed and validated risk assessment tools in the Arab region. Furthermore, similar to the findings of this study, a US-based cohort study found that FRAX (including BMD) was a superior overall predictor of hip fracture risk (AUC = 0.78 and 0.70, respectively) compared to MOF (AUC = 0.64 and 0.62). [35]

In a study in the US, the Women’s Health Initiative (WHI), which included women aged 50-64 years, found that the AUC of the OST was higher than that of FRAX. The OST in our population was slightly better than FRAX but not as high as the Women’s Initiative Study. A possible explanation could be the underrepresentation of underweight individuals in our cohort, as the OST has only three variables: age, sex, and weight. As well as by the significant interactions that were identified in this study between BMI, sex, and age, and not included in the OST [36]

Finally, this study suggests that the FRAX cutoff points of 20% for MOF and 3% for hip fracture require revision. The FRAX was found to have excellent specificity at those cutoff points, but with poor sensitivity, a finding similar to that of a systematic review and meta-analysis by Jiang et al. [37]Their review assessed the diagnostic accuracy of FRAX in predicting the 10-year risk of osteoporotic fractures, using the USA treatment thresholds for MOF and employing 20% as the 10-year fracture risk threshold for intervention. The mean sensitivity and specificity were 10.25% and 97.02%, indicating that the FRAX performed better in identifying patients who would not develop a MOF or HF within 10 years than in identifying those who would.

Our study identified eight significant risk factors. Among the available other tools assessed, the FRAX, the most commonly used in clinical practice, included eleven risk factors [5], with most of them overlapping with ours. In our model, three FRAX risk factors, viz. Rheumatoid arthritis, corticosteroid use, and alcohol use were not included due to insufficient sample size (RA and alcohol use) and lack of information (corticosteroid use).

Our cohort was similar to the US population regarding the site of fracture. Specifically, hip fractures accounted for 14% of fractures in the USA compared to 16% in our cohort. [38] An exception is vertebral fractures, which account for 6.4% of all fractures in our cohort, compared with 27% in the USA, suggesting underdiagnosis. In a study from Dubai City in the UAE, vertebral fracture prevalence accounted for 14.2% of fractures, and incidental vertebral fractures were common [39]. Underdiagnosis as an explanation is supported by Cauley et al., who concluded in a review of geographic and ethnic disparities in osteoporotic fractures that the radiographic incidence of vertebral fractures is much higher than that of hip fractures, whereas the incidence of clinical vertebral fracture is similar to that of hip fractures in most countries where data are available. [3]

Another noteworthy finding from our cohort is the high incidence of second fractures (11.9% among those with fractures) over 4.7 years, compared with a study from Korea, which reported 4.8% over 10 years [40]. This suggests that targeting patients with fractures for fracture prevention should be a priority post-discharge.

Vitamin D deficiency is potentially a modifiable risk factor. Interestingly, the association between the latest vitamin D value and fracture incidence in our study was more significant when measured at the end of follow-up, rather than at baseline. This could indicate a possible reflection of poor bone health or a surrogate indicator of poor overall healthcare-seeking behavior resulting in a lack of management of fracture risk factors, and worse lifestyle behavior choices and supplement use. Health-seeking behavior is an important risk factor, and continuity of care has been shown to reduce fracture risk. [41]

### Strengths and Limitations

This is the first study in the region to investigate fracture risk factors, and the derived model is the first of its kind. Its validation and use in clinical care will fill a significant gap in this area. This is a community-based cohort with a reasonable duration of follow-up, and outcome and covariate data were obtained by extraction of electronic medical records verified through two independent methods—chart review and direct telephone contact with patients. This dual verification likely improves the accuracy of event ascertainment and event timing, as well as of family history, behavioral factors, and medication use, partly offsetting the relatively small number of events. Applying both FRM-AD with and without BMD, with moderate predictive performance, has implications for implementing preventive strategies and planning healthcare services. Several opportunities for future research emerged from the findings of this study. First, the lack of lean body mass is a limitation in understanding the role of weight in fracture risk and in better informing management decisions. Second, having cohorts with more extended surveillance periods that start at a younger age is important, as risk factors such as vitamin D deficiency, BMI, and lifestyle factors can affect bone mineralization at a younger age and have implications later in life. Although this is the first study with a relatively large sample size, a larger cohort would have likely yielded more fractures, thereby improving our understanding of less prevalent risk factors, such as long-term medication use, rheumatoid arthritis, and thyroid diseases. Future studies are also needed to investigate the use of supplements and anti-resorptive medications prospectively.

Additionally, although fractures due to high-impact trauma and Road Traffic Collisions were excluded, the minor fractures were included, and the low incidence of hip fractures is also a limitation. Additionally, potential biases, such as self-reported data on lifestyle factors, may have existed in this study. Finally, this study lays the groundwork for planning public health interventions and informs healthcare services about gaps in screening and management that can be utilized in drafting local policies and guidelines.

Another major implication of this study is that the identified metabolic factors, such as lipids and diabetes, call for tailored interventions. Targeting bone health and metabolic risk factors at a younger age can help preserve or increase BMD, while behavioral interventions can decrease the risk of injury.

## Conclusion

This study’s findings contribute to local evidence in an understudied area and provide insights internationally. In this Emirati cohort, the locally derived FRM-AD showed moderate discrimination for incident fracture and outperformed FRAX-AD, suggesting that a locally derived model may improve fracture risk stratification. Several risk factors and predictors were identified, emphasizing priority areas for research to implement effective interventions that can enhance patients’ lives and deliver cost-effective healthcare.

## Data Availability

Data availability is restricted due to institutional policies.

## Authors’ contributions

LBK conceptualized, analyzed data, and wrote the manuscript. NN analyzed data and reviewed the manuscript. SBH, HN, MY, SA, YM, MK, YW, SS, SSH, MM, SK, AH, ES, WA, BB, AO, AK, FN, contributed to the data collection. JK reviewed the manuscript. SBH, HN, MY, and SA contributed to manuscript writing. All authors have read and approved the final manuscript.

## Acknowledgments

None.

## Consent to publish

Not Applicable.

## Availability of data and materials

Data availability is restricted due to institutional policies.

## References

1. Bassatne A, Harb H, Jaafar B, Romanos J, Ammar W, El-Hajj Fuleihan G. Disease burden of osteoporosis and other non-communicable diseases in Lebanon. Osteoporos Int. 2020;31:1769–1777.

2. GBD FC. Global, regional, and national burden of bone fractures in 204 countries and territories, 1990-2019: a systematic analysis from the Global Burden of Disease Study 2019. Lancet Healthy Longev. 2021;2:e580–e592.

3. Cauley JA, Chalhoub D, Kassem AM, Fuleihan G-H. Geographic and ethnic disparities in osteoporotic fractures. Nat Rev Endocrinol. 2014;10:338–351.

4. tool TFRAX. Fracture Risk Assessment Tool. Available from: (https://frax.shef.ac.uk/FRAX/)

5. Marques A, Ferreira RJ, Santos E, Loza E, Carmona L, da Silva JA. The accuracy of osteoporotic fracture risk prediction tools: a systematic review and meta-analysis. Ann Rheum Dis. 2015;74:1958–1967.

6. Feng JN, Zhang CG, Li BH, Zhan SY, Wang SF, Song CL. Global burden of hip fracture: The Global Burden of Disease Study. Osteoporos Int. 2024;35:41–52.

7. Michaëlsson K, Baron JA, Byberg L, Larsson SC, Melhus H, Gedeborg R. Declining hip fracture burden in Sweden 1998-2019 and consequences for projections through 2050. Sci Rep. 2024;14:706.

8. Kanis JA, Odén A, McCloskey EV, Johansson H, Wahl DA, Cooper C et al. A systematic review of hip fracture incidence and probability of fracture worldwide. Osteoporos Int. 2012;23:2239–2256.

9. Warriner AH, Patkar NM, Curtis JR, Delzell E, Gary L, Kilgore M et al. Which fractures are most attributable to osteoporosis. J Clin Epidemiol. 2011;64:46–53.

10. Cauley JA, Thompson DE, Ensrud KC, Scott JC, Black D. Risk of mortality following clinical fractures. Osteoporos Int. 2000;11:556–561.

11. Bao Y, Xu Y, Li Z, Wu Q. Racial and ethnic difference in the risk of fractures in the United States: a systematic review and meta-analysis. Sci Rep. 2023;13:9481.

12. Chakhtoura M, Dagher H, Sharara S, Ajjour S, Chamoun N, Cauley J et al. Systematic review of major osteoporotic fracture to hip fracture incidence rate ratios worldwide: implications for Fracture Risk Assessment Tool (FRAX)-derived estimates. J Bone Miner Res. 2021;36:1942–1956.

13. Dawod MS, Alisi MS, Saber YO, Abdel-Hay QA, Al-Aktam BM, Alfaouri Y et al. Characteristics of Elderly Hip Fracture Patients in Jordan: A Multicenter Epidemiological Study. Int J Gen Med. 2022;15:6591–6598.

14. Abdulla N, Alsaed OS, Lutf A, Alam F, Abdulmomen I, Al Emadi S et al. Epidemiology of hip fracture in Qatar and development of a country specific FRAX model. Arch Osteoporos. 2022;17:49.

15. Barake M, El Eid R, Ajjour S, Chakhtoura M, Meho L, Mahmoud T et al. Osteoporotic hip and vertebral fractures in the Arab region: a systematic review. Osteoporos Int. 2021;32:1499–1515.

16. Koh LK, Sedrine WB, Torralba TP, Kung A, Fujiwara S, Chan SP et al. A simple tool to identify asian women at increased risk of osteoporosis. Osteoporos Int. 2001;12:699–705.

17. Latifa Baynouna Alketbi, Shahra Bin Hraiz, Hessa AlNeyadi, Mahra Alyahyaei, Shamsa AlAlawi, Yasmin Mohamed, Maitha Alkaabi, Juma AlKaabi, Sara Al Shukri, Shamsa Alshamsi, Mohammad Moussa, Sanaa Al Kalbani, Amal Hantash, Ekram Saeed, Waad Alantali, Besent Elbeheiry, Yahya Alwaqfi, AbdulRahman Omara, Amal Al Khouri, Nico Nagelkerke. Osteoporosis prevalence, incidence, and predictors in a population-based cohort study in Abu Dhabi. Unpublished. 2025

18. US PSTF, Nicholson WK, Silverstein M, Wong JB, Chelmow D, Coker TR et al. Screening for Osteoporosis to Prevent Fractures: US Preventive Services Task Force Recommendation Statement. JAMA. 2025;333:498–508.

19. Agrawal AC, Garg AK. Epidemiology of Osteoporosis. Indian J Orthop. 2023;57:45–48.

20. Jung Y, Ko Y, Kim HY, Ha YC, Lee YK, Kim TY et al. Gender differences in antiosteoporosis drug treatment after osteoporotic fractures. J Bone Miner Metab. 2019;37:134–141.

21. Nielson CM, Marshall LM, Adams AL, LeBlanc ES, Cawthon PM, Ensrud K et al. BMI and fracture risk in older men: the osteoporotic fractures in men study (MrOS). J Bone Miner Res. 2011;26:496–502.

22. Xiang BY, Huang W, Zhou GQ, Hu N, Chen H, Chen C. Body mass index and the risk of low bone mass-related fractures in women compared with men: A PRISMA-compliant meta-analysis of prospective cohort studies. Medicine (Baltimore). 2017;96:e5290.

23. Gandham A, Zengin A, Bonham MP, Winzenberg T, Balogun S, Wu F et al. Incidence and predictors of fractures in older adults with and without obesity defined by body mass index versus body fat percentage. Bone. 2020;140:115546.

24. Chan MY, Frost SA, Center JR, Eisman JA, Nguyen TV. Relationship between body mass index and fracture risk is mediated by bone mineral density. J Bone Miner Res. 2014;29:2327–2335.

25. Turcotte AF, Jean S, Morin SN, Mac-Way F, Gagnon C. Relationships between Obesity and Incidence of Fractures in a Middle-Aged Population: A Study from the CARTaGENE Cohort. JBMR Plus. 2023;7:e10730.

26. Shiomoto K, Babazono A, Harano Y, Fujita T, Jiang P, Kim SA et al. Effect of body mass index on vertebral and hip fractures in older people and differences according to sex: a retrospective Japanese cohort study. BMJ Open. 2021;11:e049157.

27. Yoon H, Sung E, Kang JH, Kim CH, Shin H, Yoo E et al. Association between body fat and bone mineral density in Korean adults: a cohort study. Sci Rep. 2023;13:17462.

28. Ho-Pham LT, Nguyen UD, Nguyen TV. Association between lean mass, fat mass, and bone mineral density: a meta-analysis. J Clin Endocrinol Metab. 2014;99:30–38.

29. Parhami F, Garfinkel A, Demer LL. Role of lipids in osteoporosis. Arterioscler Thromb Vasc Biol. 2000;20:2346–2348.

30. Shao L, Luo S, Zhao Z. Lipid metabolites are associated with the risk of osteoporotic fractures. Sci Rep. 2024;14:19245.

31. Skjødt MK, Möller S, Hyldig N, Clausen A, Bliddal M, Søndergaard J et al. Validation of the Fracture Risk Evaluation Model (FREM) in predicting major osteoporotic fractures and hip fractures using administrative health data. Bone. 2021;147:115934.

32. Dautzenberg L, Beglinger S, Tsokani S, Zevgiti S, Raijmann RCMA, Rodondi N et al. Interventions for preventing falls and fall-related fractures in community-dwelling older adults: A systematic review and network meta-analysis. J Am Geriatr Soc. 2021;69:2973–2984.

33. Wu CH, Tu ST, Chang YF, Chan DC, Chien JT, Lin CH et al. Fracture liaison services improve outcomes of patients with osteoporosis-related fractures: A systematic literature review and meta-analysis. Bone. 2018;111:92–100.

34. Fink HA, Butler ME, Claussen AM, Collins ES, Krohn KM, Taylor BC et al. Performance of Fracture Risk Assessment Tools by Race and Ethnicity: A Systematic Review for the ASBMR Task Force on Clinical Algorithms for Fracture Risk. J Bone Miner Res. 2023;38:1731–1741.

35. Hillier TA, Cauley JA, Rizzo JH, Pedula KL, Ensrud KE, Bauer DC et al. WHO absolute fracture risk models (FRAX): do clinical risk factors improve fracture prediction in older women without osteoporosis. J Bone Miner Res. 2011;26:1774–1782.

36. Crandall CJ, Larson JC, Schousboe JT, Manson JE, Watts NB, Robbins JA et al. Race and Ethnicity and Fracture Prediction Among Younger Postmenopausal Women in the Women’s Health Initiative Study. JAMA Intern Med. 2023;183:696–704.

37. Jiang X, Gruner M, Trémollieres F, Pluskiewicz W, Sornay-Rendu E, Adamczyk P et al. Diagnostic accuracy of FRAX in predicting the 10-year risk of osteoporotic fractures using the USA treatment thresholds: A systematic review and meta-analysis. Bone. 2017;99:20–25.

38. Burge R, Dawson-Hughes B, Solomon DH, Wong JB, King A, Tosteson A. Incidence and economic burden of osteoporosis-related fractures in the United States, 2005-2025. J Bone Miner Res. 2007;22:465–475.

39. Majumdar SR, Kim N, Colman I, Chahal AM, Raymond G, Jen H et al. Incidental vertebral fractures discovered with chest radiography in the emergency department: prevalence, recognition, and osteoporosis management in a cohort of elderly patients. Arch Intern Med. 2005;165:905–909.

40. Ha YC, Park YG, Nam KW, Kim SR. Trend in hip fracture incidence and mortality in Korea: a prospective cohort study from 2002 to 2011. J Korean Med Sci. 2015;30:483–488.

41. Kim SH, Kim H, Jeong SH, Jang SY, Park EC. Impact of continuity of care on risk for major osteoporotic fracture in patients with new onset rheumatoid arthritis. Sci Rep. 2022;12:10189.

