## Supplementary figures and images for "Incidence of Fractures and Development of the Fracture Risk Model (FRM) in a Population-Based Cohort Study in Abu Dhabi"

### Appendix 1

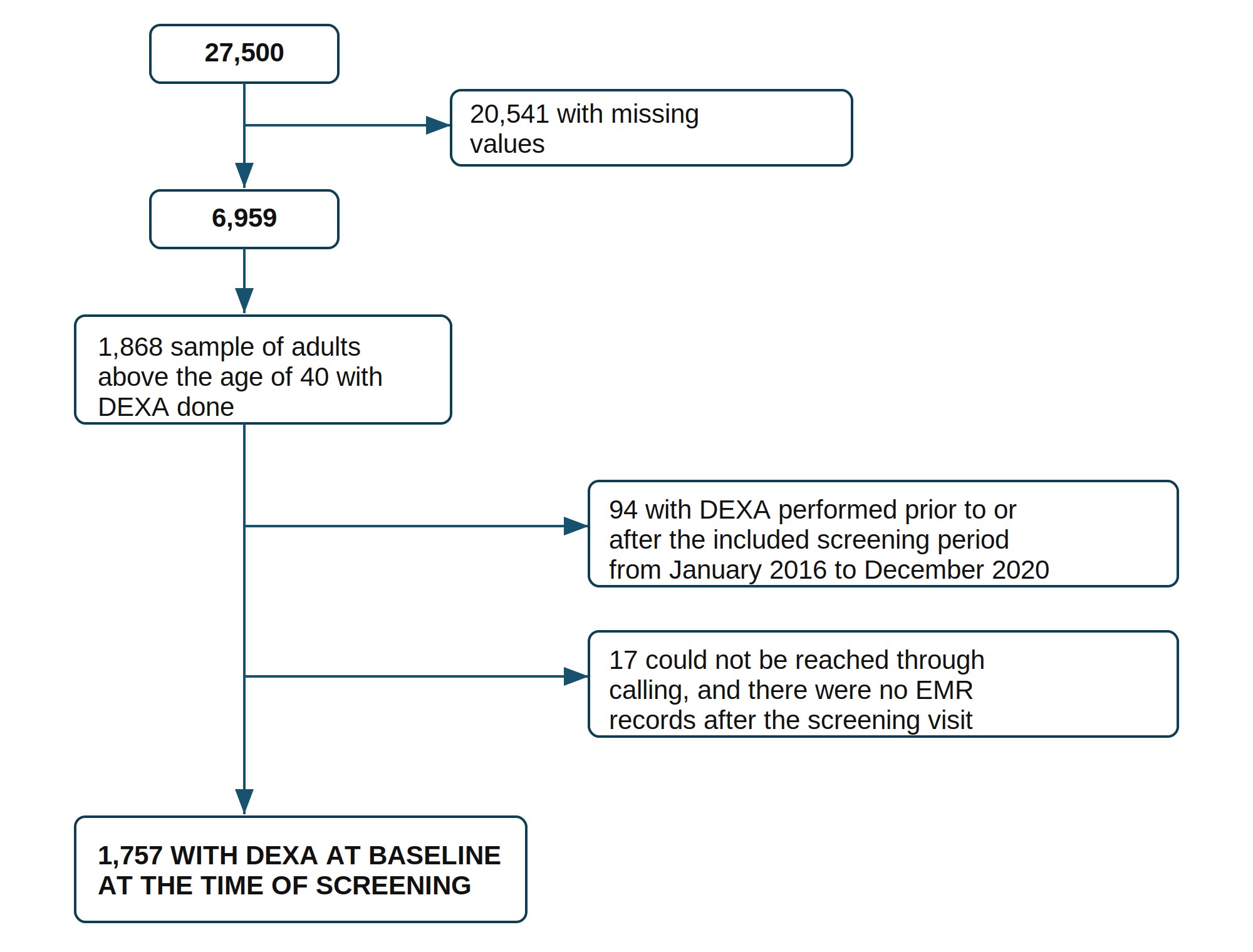

### Appendix 2

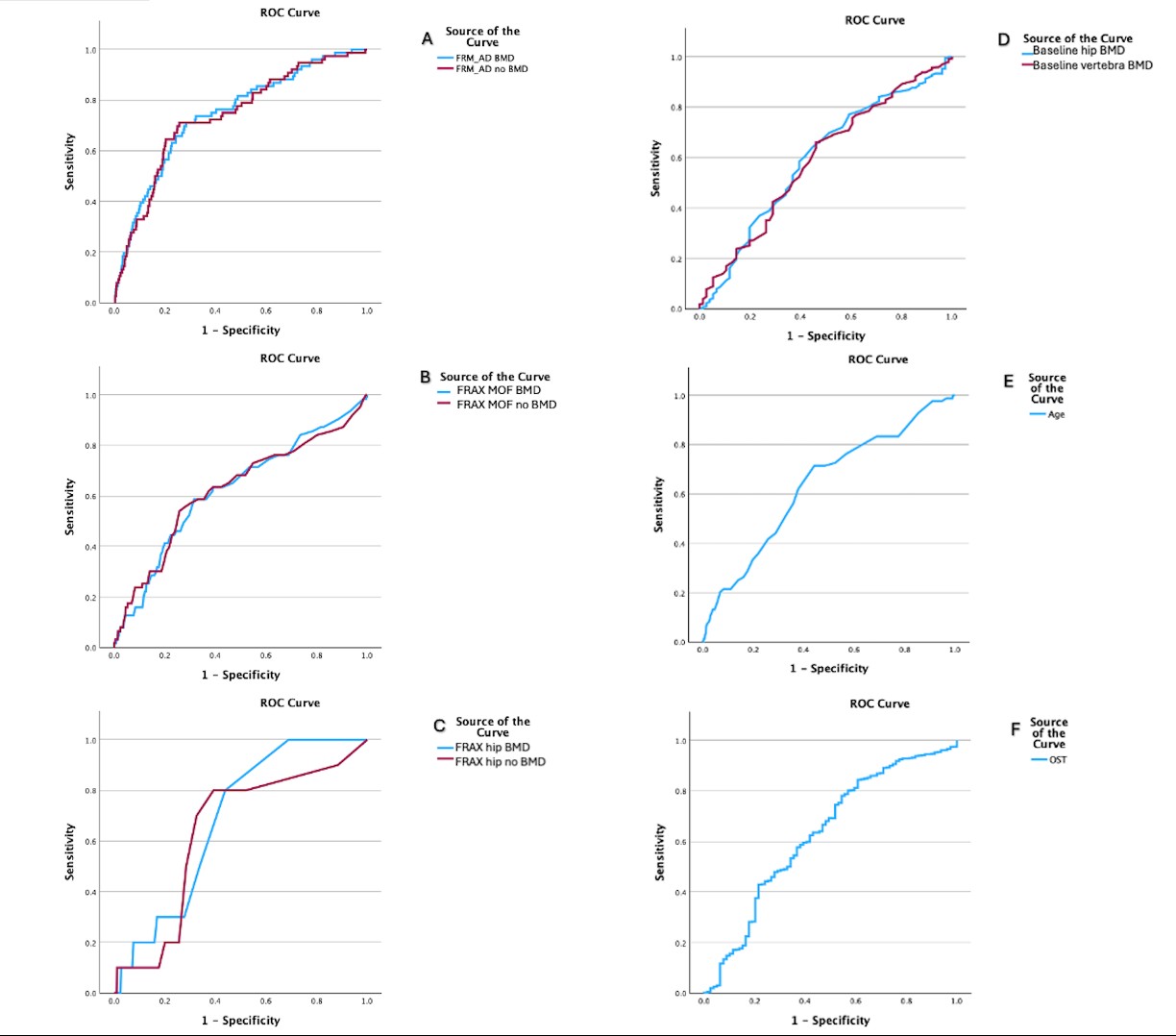

### Appendix 3

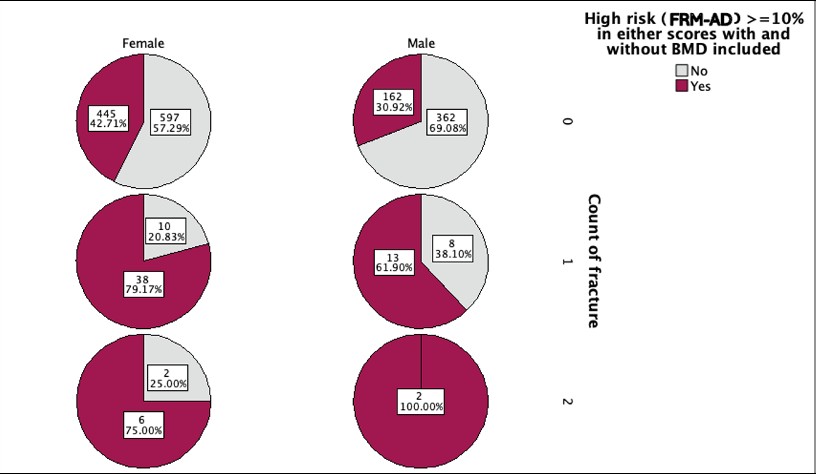

### Appendix 4

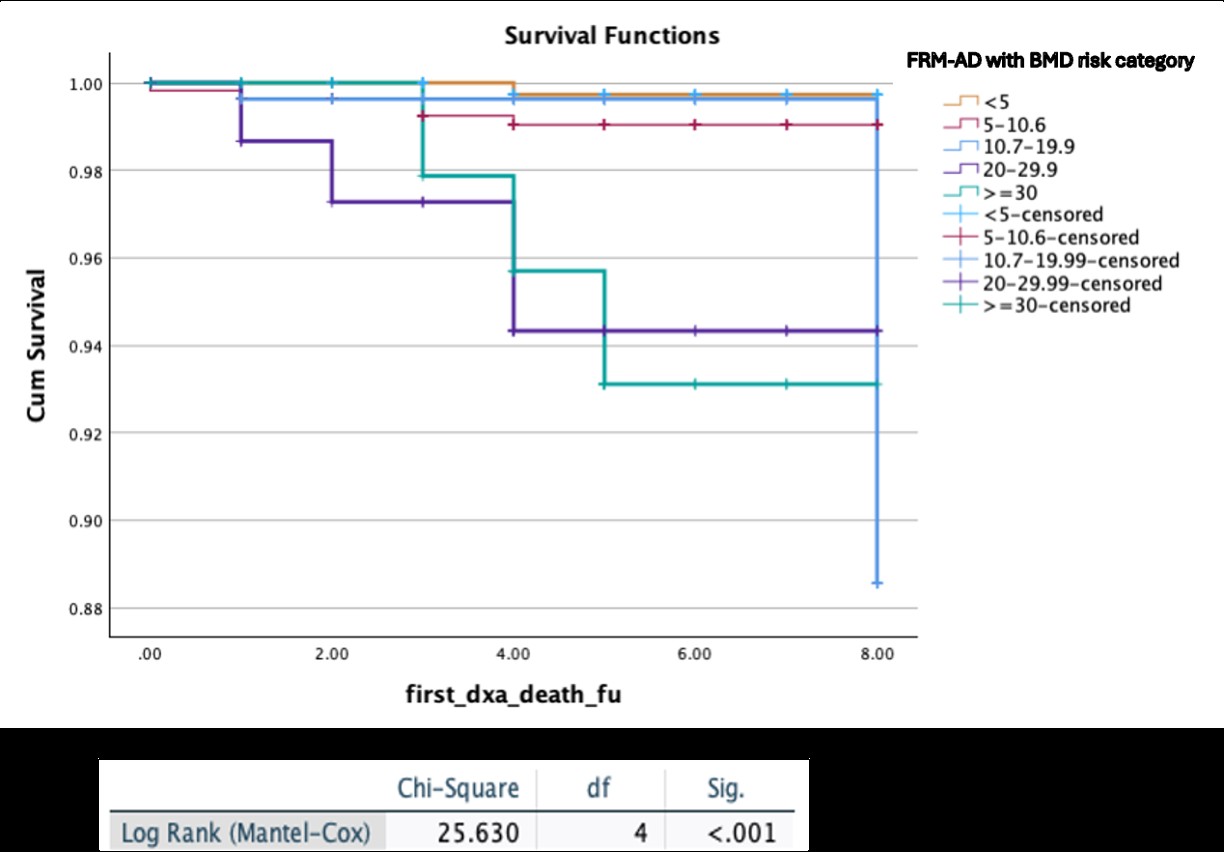
