## Appendix 5 for "Incidence of Fractures and Development of the Fracture Risk Model (FRM) in a Population-Based Cohort Study in Abu Dhabi"

Appendix 5: Sensitivity, Specificity, Negative Predictive Value (NPV), and Positive Predictive Value (PPV) of the FRM-AD for identifying fractures. A, with no BMD included; B, with BMD included.

A. No BMD

| cutoff | Sensitivity | Specificity | NPV | PPV |
| --- | --- | --- | --- | --- |
| 5 | 94.9 | 24.5 | 99.0 | 6.0 |
| 10 | 69.6 | 68.7 | 97.8 | 10.1 |
| 15 | 32.9 | 87.0 | 96.3 | 11.4 |
| 20 | 25.3 | 93.4 | 96.1 | 16.3 |
| 30 | 10.1 | 97.9 | 95.6 | 19.5 |

B. With BMD

| cutoff | Sensitivity | Specificity | NPV | PPV |
| --- | --- | --- | --- | --- |
| 5 | 88.2 | 30.6 | 98.0 | 6.3 |
| 10 | 71.1 | 68.9 | 97.8 | 10.8 |
| 15 | 46.1 | 84.4 | 96.7 | 13.5 |
| 20 | 32.9 | 91.4 | 96.3 | 16.9 |
| 30 | 15.8 | 96.7 | 95.6 | 20.3 |
